# Implicit Bias And Unintentional Harm In Vascular Care: The Case For Intervention

**DOI:** 10.1101/2024.02.13.24302798

**Authors:** Corey A. Kalbaugh, Erika T. Beidelman, Kerry A. Howard, Brian Witrick, Ashley Clark, Katharine L. McGinigle, Samantha Minc, Olamide Alabi, Caitlin W. Hicks, Andrew A. Gonzalez, Crystal W. Cené, Samuel Cykert

## Abstract

**Introduction:** Implicit (or unconscious) bias may influence physician treatment decisions and contribute to Black-White health disparities. While implicit bias has been linked with low quality care via clinical vignettes, some worry that these studies are not representative of the ‘real world.’ There is limited data that has attempted to link implicit bias with actual care delivery and outcomes. We sought to understand if implicit bias is associated with potentially harmful surgical treatment selection in a cohort of patients with peripheral artery disease (PAD)-related claudication undergoing below-knee lower extremity revascularization as captured in a ‘real world’ procedural registry.

**Methods:** We invited vascular specialists from the Vascular Quality Initiative (VQI) to take the race Implicit Association Test (IAT). The IAT asks participants to associate images of Black and White Americans with either positive or negative attributes. Based on reaction time differences across sequential tests, participants were grouped into race-based implicit bias categories: pro-White bias, no bias, or pro-Black bias. Our provider-level implicit bias results were linked to patient-level registry data of peripheral revascularization procedures performed for claudication. We measured the adjusted odds of performance of below knee procedures by specialist implicit bias and patient race via mixed effects logistic regression models. We assessed implicit bias as a moderator of the association of below-knee procedures and patient race with 1-yr amputation.

**Results:** 218 vascular specialists in the United States completed the IAT and 157 (72%) had a pro-White bias. Black patients treated by a physician with pro-White bias had a 74% increase in the odds of receiving a below-knee procedure compared to the total sample (aOR: 1.74, 95% CI: 1.33-2.15). When treated by a specialist with pro-White bias, Black patients had 3 times the odds of 1-yr amputation – regardless of anatomic location treated – compared to White patients (aOR: 3.04, 95% CI: 1.68-5.51). Conversely, Black patients treated by a specialist with no bias had similar odds of a below-knee procedure (aOR: 0.99, 95% CI: 0.67-1.30) and 1-year amputation (aOR: 1.31, 95% CI: 0.35-4.96) as the full patient sample.

**Conclusions:** Implicit bias is associated with potentially harmful below-knee procedures for Black patients and contributes to Black-White outcome disparities in the United States. These results suggest the need for system-level interventions that transparently identify and warn of procedures not aligned with best practices to reduce the negative effect of implicit bias.

**Clinical Perspective:** *What is new?:*

- Pro-White bias is associated with low-value care for Black patients.
- Black patients treated by physicians with pro-White implicit preferences also had worse 1-year amputation rates.
- We found little race-based variation in treatment decisions for below-knee revascularization procedures among physicians with no implicit bias as measured by the IAT.

*What are the clinical implications?:*

- Physicians who treat vascular diseases consider their own practices and where they may be falling short of standards, particularly for their Black patients.
- Health care leaders must begin to evaluate how and why their own health care systems make it possible for practitioners’ unconscious biases to negatively impact care.
- Policy changes that enhance payment for evidence-based procedures and reduce payment for potentially harmful procedures should be considered.

## INTRODUCTION

Nearly ten million people in the United States have claudication, a subtype of peripheral artery disease (PAD) characterized by exercise-induced pain or fatigue in the lower extremities.^1^ Claudication can cause severe lifestyle limitations and may progress to premature functional impairment and death.^2^ More than a decade’s worth of research has shown worse outcomes for Black patients with claudication compared to White patients with claudication.^3^ The reasons behind the persistence of these inequities remain poorly described. We have examined how the clinical management of patients with claudication differs by patient race and the role this differential management plays in Black-White health inequities. Black patients are less likely to be offered a limb salvage attempt prior to amputation in situations where limb vitality is threatened.^4-6^ Black patients are also less likely to be offered guideline-directed medical therapy than their White counterparts.^7^ Additionally, Black individuals residing in low-income counties are significantly more likely to undergo early and potentially harmful below-knee peripheral revascularizations for claudication compared to similar White individuals^8,9^. Black individuals with PAD in non-distressed communities have a higher lifetime incidence of amputation compared to White individuals in distressed communities.^10^ A better understanding of the mechanisms that underlie these racial inequities in both potentially harmful revascularizations and limb loss is necessary to ultimately provide better care and improved outcomes.

So *why* do Black patients receive different and suboptimal care compared to White patients even when controlling for location, insurance, socioeconomic status (SES), and risk factor profiles? One potential mechanism contributing to care inequities is implicit bias, defined as an unconscious bias that may influence behavior, cognitive processes, and decision-making.^11^ As described in several recent systematic reviews, health care providers have implicit biases and that may impact the delivery of health care in a variety of settings.^12-14^ For example, physician bias and stereotyping may have an influence on who receives the “best” health care interventions and who receives harmful care even if unintentional.^15,16^ However, while implicit bias has been linked with low quality care in the setting of clinical vignettes,^17^ these studies have been critiqued as lacking the stressors present in a “real-life” clinical environment.^12^ Conversely, implicit bias tests are able to more directly measure the unconscious association through reaction time. There is a paucity of data exploring the relationship between implicit bias among physicians with actual care delivery and related outcomes.

The purpose of our study was to examine if physician implicit bias is associated with the provision of low-value care for Black or White patients with claudication, among a sample of vascular specialists using the implicit association test (IAT)^18^. We defined low-value care as the performance of below-knee peripheral revascularization procedures. We also assessed the association between below-knee procedures and patient race on the odds of a 1-year amputation, using physician implicit bias as a moderator.

## METHODS

### Study Sample and Source Population

The Society for Vascular Surgery (SVS) Vascular Quality Initiative (VQI) is a vascular procedure-based registry^19^ intended to document and improve the quality of care delivered to patients with vascular diseases. Clinicians that participate in the delivery of vascular care in the United States are eligible to participate in the VQI. As of January 2023, the VQI includes 960 sites. The VQI has 18 regional quality groups within four regions: South (5), Northeast (4), Midwest (4), and West (5). Centers include academic medical centers, teaching hospitals, community hospitals, and private practices. We invited all vascular specialist VQI members (N=2,512) to take the race Implicit Association Test (IAT) to categorize their implicit bias level. We then linked the IAT to procedure-level data from the VQI registry. The registry contains information on patient demographics, comorbid conditions, imaging studies, medication usage, peri-procedural details, and in-hospital and thirty-day outcomes. Patients also are expected to complete a one-year follow-up visit for any procedure covered by the VQI and outcome data at one-year are included in the registry. The Social Security Death Index is linked to the VQI registry to ascertain deaths. We restricted the procedure-level dataset to observations linked to only White or Black patients as the race IAT only measures implicit bias in relation to White or Black individuals.

### Exposure: Implicit Bias

The IAT is an online assessment that takes 10 to 15 minutes to complete and asks participants to associate images of White and Black individuals with either positive or negative attributes. It is the most widely used test for studying unconscious bias in scientific research with established acceptable levels of reliability and validity.^18,20^ Based on the differences in reaction times across sequential tests, participants were grouped into seven implicit bias categories based on racial preference: Strong pro-White bias, moderate pro-White bias, slight pro-White bias, no bias, slight pro-Black bias, moderate pro-Black bias, and strong pro-Black bias. We then further reduced this categorical variable into the following three levels due to low within-category sample sizes: pro-White bias, no bias, and pro-Black bias.

### Primary Outcome: Risk>Benefit Procedures

To test the association between physician implicit bias and procedure performance, we assessed the outcome of below the knee procedures for claudication because these have been identified as carrying high risk with questionable benefit.^21,22^ We defined below-knee as arteries inclusive of the below-knee popliteal and arteries more distal (tibial, pedal, etc). Because these procedures often lead to major adverse vascular events, we used this as our primary outcome.^23^ Performance of a below-knee procedure was represented by a binary indicator variable with ‘1’ indicating the procedure was performed and ‘0’ indicating it was not.

### Secondary Outcome: 1-Year Amputation

The VQI registry longitudinally follows and records patient outcomes following each procedure including instances of major amputation. Therefore, we examined the rates of major amputation within 1-year of recorded procedure. We used the date of the procedure and the date of amputation to determine amputations that occurred within the year following the procedure. Minor amputations were not included in analyses.

### Covariates

Both physician-level and procedure-level covariates were included in our analysis to describe the sample and account for confounding. Physician-level characteristics included age (years), gender (male or female), specialty (cardiothoracic surgery, interventional cardiology, interventional radiology, or vascular surgery), years of experience (0-2 years, 3-5 years, 6-10 years, 11-15 years, 16-20 years, 21-25 years, or 26+ years), race/ethnicity (non-Hispanic white, non-Hispanic black, non-Hispanic other, or Hispanic), and region (Midwest, Northeast, South, or West). Procedure-level characteristics included year of procedure, anatomic location of the intervention, type of intervention (endovascular or open bypass), patient age, gender, diabetes status, current smoking status (yes or no), body-mass index, and patient race (non-Hispanic white or non-Hispanic black).

### Statistical Analysis

To reduce the potential effect of differences in the characteristics of physicians from the SVS VQI registry who participated compared to the full registry, data were weighted. First, we cross-classified physicians into categories (or “weighting cells”) based on the following three variables: US geographic region, age category, and sex, due to differences in the likelihood of participating in the study (for example, physicians who identified as female were more likely to participate). Then, a weight defined as the number of physicians on the registry divided by the number of physicians who participated in the study was applied within each “weighting cell” (e.g., male physicians 30-44 years of age from the Northeast region of the United States) to “weight up” physicians from demographic groups who were underrepresented in the data. Analyses presented here account for weighting.

We then performed univariate descriptive analyses to characterize the physicians and procedures included in our analytical sample. We compared the distribution of physician-level sociodemographic characteristics across the three identified implicit bias categories. To answer our research questions, we restricted our dataset to procedures linked to patients that presented with claudication. To assess the association between physician implicit bias and the performance of below knee procedures among patients with claudication, we fit mixed effects logistic regression models. With these models we estimated the odds of a below knee procedure being performed associated within each physician implicit bias category. All models included a random intercept for the physician to account for correlation between multiple procedures performed by the same physician. Additionally, all models included an interaction between physician implicit bias category and patient race. We included this interaction term as we anticipated that the association between implicit bias and procedure performance would vary based on patient race/ethnicity. We also report interaction contrasts for the adjusted model. Ultimately, we reduced our exposure categorization to two levels (No Bias/pro-Black bias versus Pro-White Bias) due to non-convergence of models that included the pro-Black bias group. This is due to the low sample size within the pro-Black bias exposure group.

We also assessed the association between both below-knee procedures and patient race on the odds of a 1-year amputation, using physician implicit bias as a moderator. We estimated all associations via mixed effects models with a random intercept for the physician to account for the correlation between procedures performed by the same physician. To assess moderation by physician implicit bias, we stratified our analysis by the three levels of implicit bias. Due to insufficient sample size, we could not fit a model for the Black implicit bias preference category in the model assessing the association between the performance of below knee procedures and the odds of a 1-year amputation. All statistical analyses were performed in RStudio version 4.1.1 and the mixed effects models were fit using the lme4 package.^24^

## RESULTS

### Sample Vascular Specialists Characteristics

We had 218 vascular specialists that met inclusion criteria of 1) completing the entire IAT survey and 2) were able to be linked to procedural-level data in the VQI registries. The majority of physician participants were vascular surgeons (91%), non-Hispanic White (66%), and male (74%) (*Table 1*). Non-white physicians included non-Hispanic Black (n=10), Hispanic (n=9), non-Hispanic Other (n=55) (including physicians identifying as multiracial, Asian, American Indian/Alaska Native, Native Hawaiian, Other Pacific Islander, and Other). The median age of participants was 46 years and median years of practice was 6-10 years.

**Table 1.**
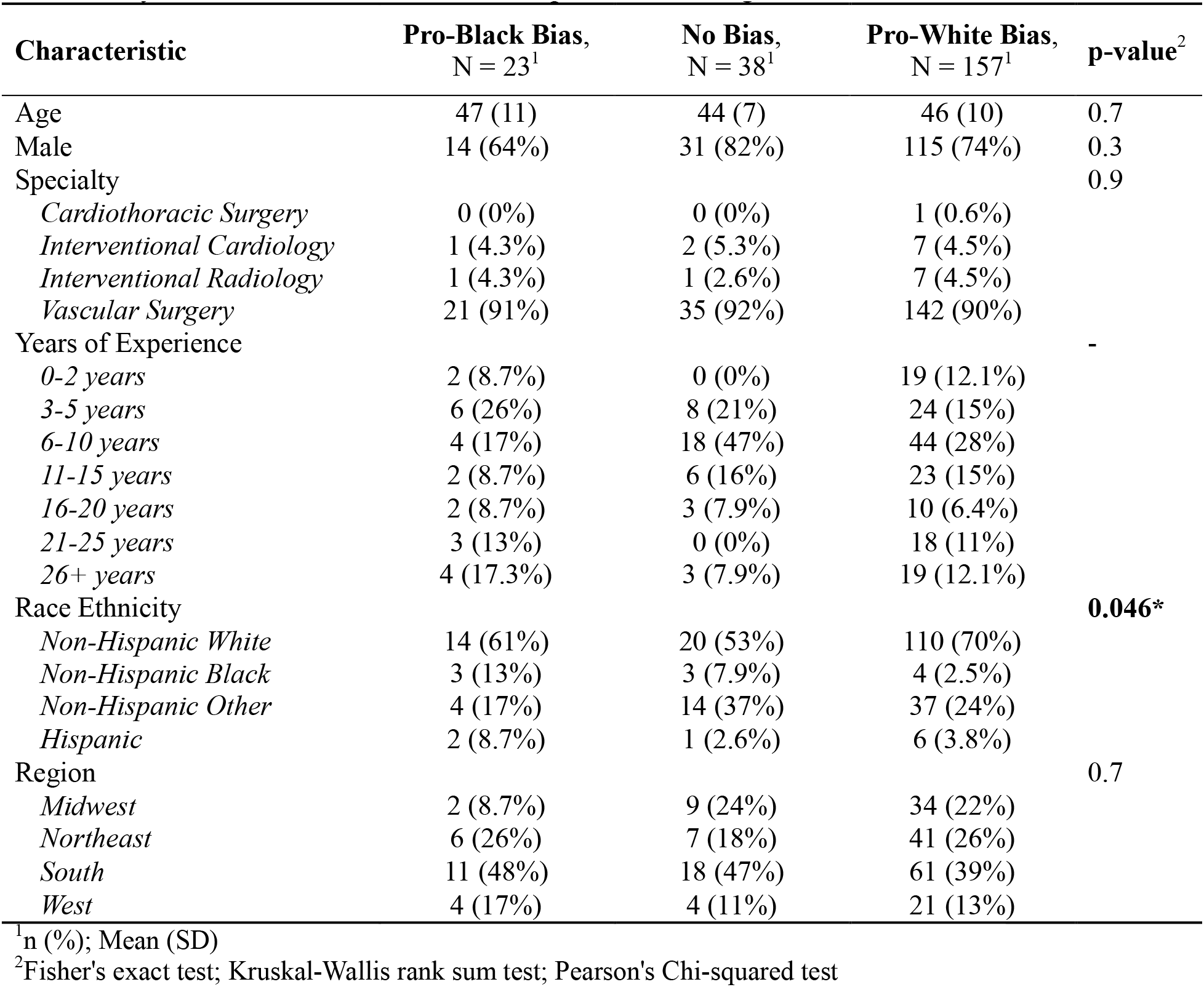
Physician Characteristics Across Implicit Bias Categories.

### Sample Patient Characteristics

Our 218 vascular specialists were linkable to 6,588 patients on whom they performed a peripheral revascularization procedure for claudication. The majority of the procedures were endovascular (n=4947; 75%). There were 880 below-knee procedures and Black patients had a higher prevalence of these procedures than White patients (19% v 13%). Compared to White patients, Black patients were younger, more likely to be female, and had a higher prevalence of current smoking status and diabetes (*Table 2*).

**Table 2.** Patient Characteristics Overall and by Patient Race.

| **Characteristic** | **Overall**<br>N = 6,588<sup>1</sup> | **White**<br>N = 5,399<sup>1</sup> | **Black**<br>N = 1,189<sup>1</sup> | **p-value**<sup>2</sup> |
| --- | --- | --- | --- | --- |
| Age | 67 (10) | 67 (10) | 65 (10) | <0.001 |
| BMI | 28.4 (6.0) | 28.4 (5.9) | 28.3 (6.4) | 0.4 |
| Male | 4,335 (66%) | 3,687 (68%) | 648 (54%) | <0.001 |
| Current Smoker | 2,635 (40%) | 2,110 (39%) | 525 (44%) | 0.001 |
| Diabetes | 2,826 (43%) | 2,222 (41%) | 604 (51%) | <0.001 |
| Below Knee | 880 (14%) | 661 (13%) | 219 (19%) | <0.001 |

<sup>1</sup>Mean(SD); n (%)
<sup>2</sup>Wilcoxon rank sum test; Pearson's Chi-squared test

### Physician Implicit Bias

Based on the results of the IAT, we found that 72% of vascular specialists had a pro-White bias, 17% had no specific preference for Black or White individuals, and 11% had a pro-Black bias. Of the 157 specialists with a pro-White bias, 39 were found to have a slight bias, 70 had a moderate bias, and 48 had a strong bias for White compared to Black individuals. Of the 23 specialists with a pro-Black bias, 10 were found to have a slight bias, 10 had a moderate bias, and 3 had a strong bias for Black compared to White individuals.

### Physician Implicit Bias and Below-Knee Procedures

From an adjusted model, we found a significant interaction between physician pro-White bias and patient race on the odds of a below-knee procedure among patients with claudication (aOR: 1.59, 95% CI: 1.02-2.49) (see *Table 3)*. Black patients treated by a physician with pro-White bias had a 74% increase in the odds of receiving a below-knee procedure compared to the total sample (aOR: 1.74, 95% CI: 1.33-2.15) (see *Figure 1*). In contrast, White patients treated by both physicians with pro-White bias or no bias had a 19% and 28% reduced odds of a below-knee procedure, respectively, compared to the sample (Physician with pro-White bias aOR: 0.81, 95% CI: 0.64-0.97; Physician with no bias aOR: 0.72, 95% CI: 0.56-0.89).

**Table 3.** Odds of a Below-Knee Procedure Among Patients with Claudication.

| *Predictors* | **Unadjusted** |  |  | **Adjusted** |  |  |
| --- | --- | --- | --- | --- | --- | --- |
|  | *Odds Ratios* | *CI* | *p* | *Odds Ratios* | *CI* | *p* |
| (Intercept) | 0.13 | 0.09 – 0.18 | **<0.001** | 0.16 | 0.07 – 0.34 | **<0.001** |
| No Bias |  | *Reference* |  |  | *Reference* |  |
| Pro-White Bias | 1.12 | 0.78 – 1.60 | 0.537 | 1.12 | 0.77 – 1.63 | 0.546 |
| White Patient Race |  | *Reference* |  |  | *Reference* |  |
| Black Patient Race | 1.30 | 0.88 – 1.91 | 0.185 | 1.36 | 0.92 – 2.01 | 0.129 |
| No Bias*Black Patient Race |  | *Reference* |  |  | *Reference* |  |
| Pro-White Bias*Black Patient Race | 1.42 | 0.91 – 2.19 | 0.119 | 1.59 | 1.02 – 2.49 | **0.040** |

Adjusted for physician race, year, patient gender, patient smoking status, diabetes status, and patient age

**Figure 1.**
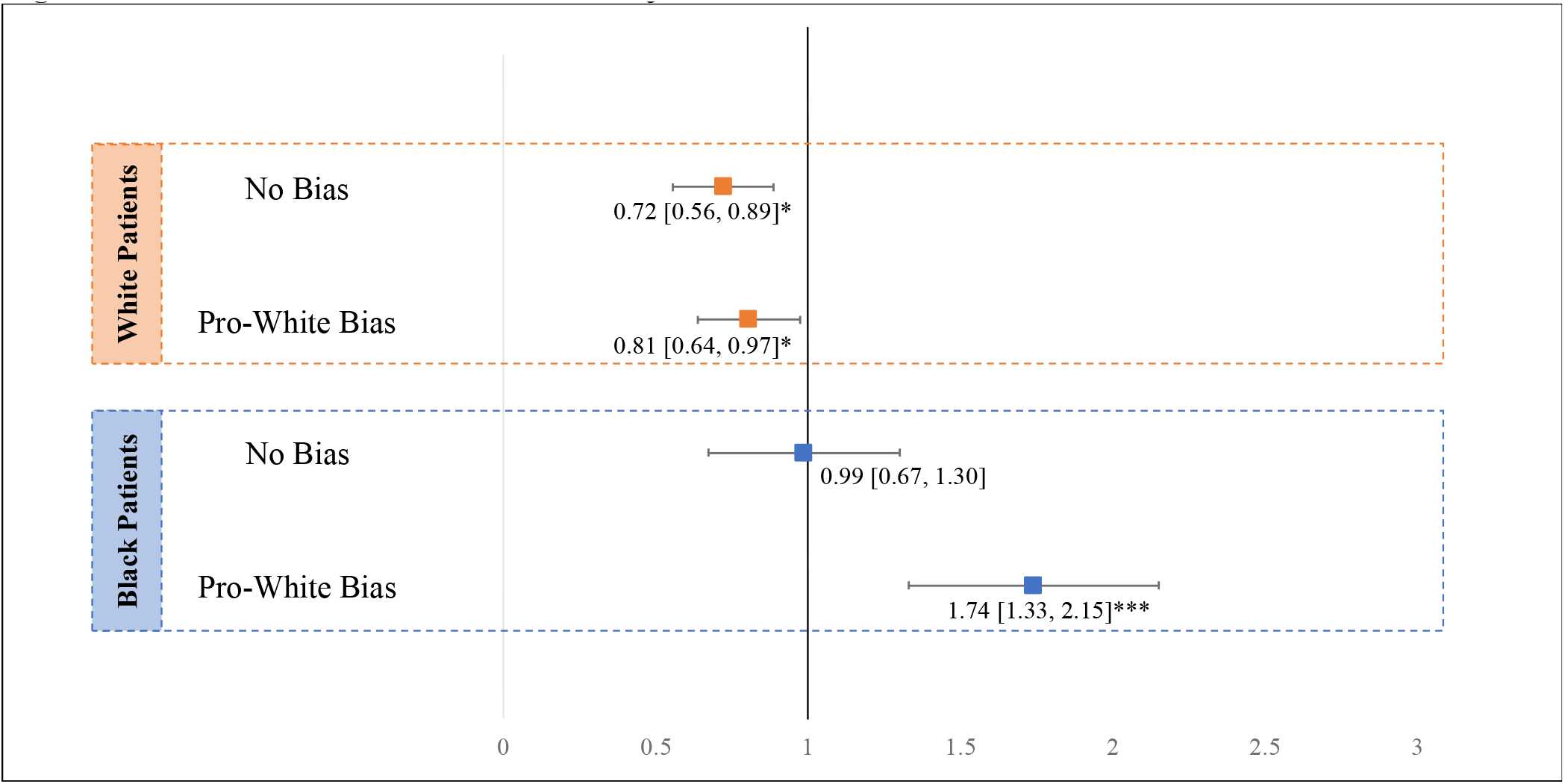
Interaction Contrasts from the Adjusted Below-Knee Procedure Model

**Figure 2.**
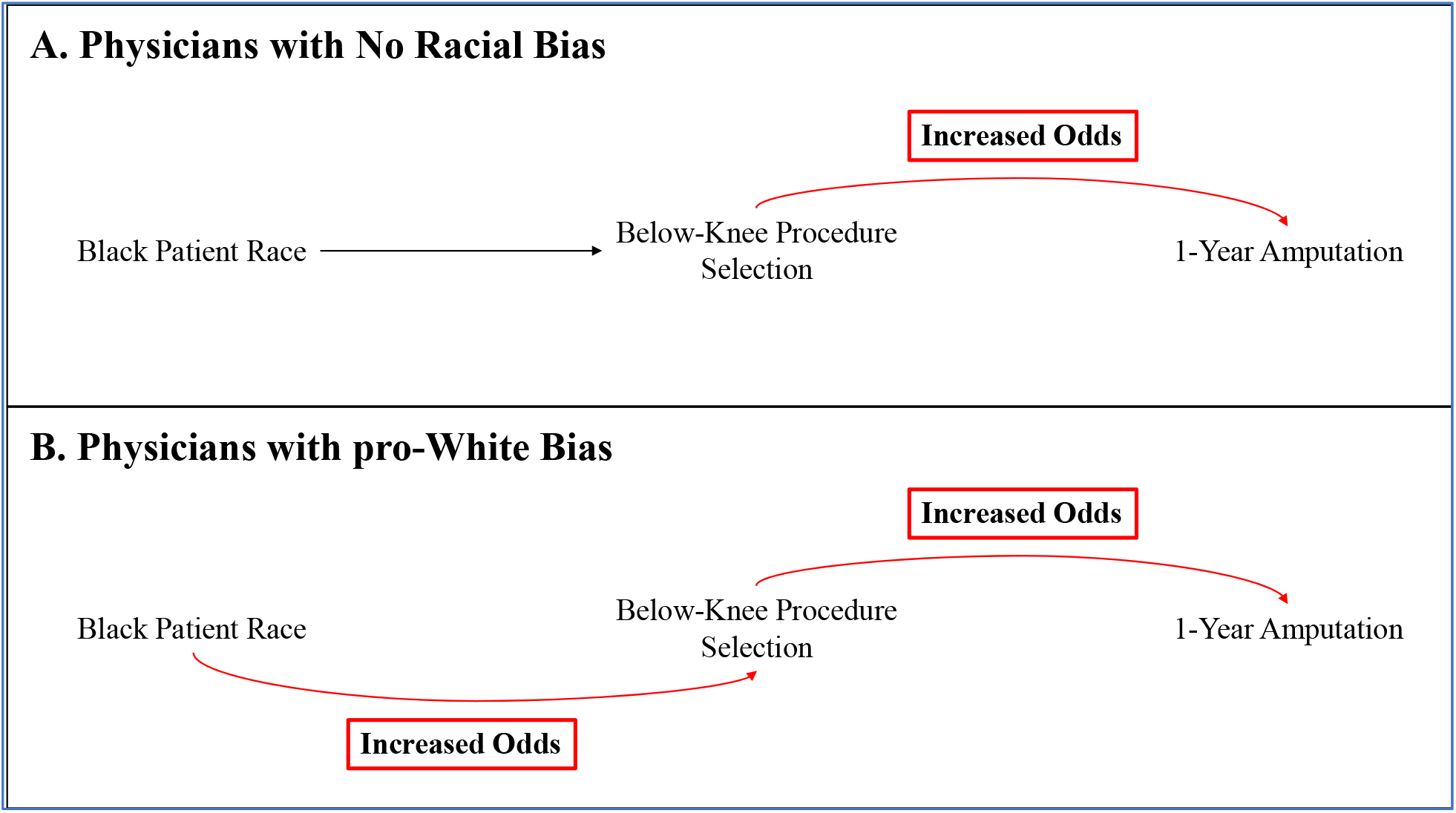
Conceptual Model Illustrating the Moderating Effect of Physician Implicit Bias on Amputation Outcomes Among Black Patients with Claudication

### 1-Year Incidence of Major Amputation

The overall one-year incidence of major amputation was 1.10 (0.77,1.30) in our overall population of patients treated for claudication. Incidence of amputation was higher among Black patients compared to White patients (1.94% v 0.81%; aHR: 2.14, 95% CI: 1.23, 3.72).

### 1-Year Amputation and Physician Implicit Bias as a Moderator

Among patients treated by a physician with no racial bias, patient race was not associated with the odds of 1-year amputation (aOR: 1.31, 95% CI: 0.35-4.96) (see *Table 4*). Among patients treated by physicians with a pro-White bias, Black patients had 3.3 times the odds of experiencing an amputation within 1-year of the procedure compared to White patients (aOR: 3.26, 95% CI: 1.80-5.90). Among patients treated by a physician with no racial bias, patients receiving a below-knee procedure had 4.5 times the odds of experiencing an amputation within 1-year compared to above-knee procedures (aOR: 4.47, 95% CI: 1.63-12.21) (see *Table 5*).

**Table 4.** Odds of 1-Year Amputation Associated with Patient Race Stratified by Implicit Bias Category, among patients with claudication.

| *Predictors* | **No Bias** |  |  | **Pro-White Bias** |  |  |
| --- | --- | --- | --- | --- | --- | --- |
|  | *Odds Ratios* | *CI* | *p* | *Odds Ratios* | *CI* | *p* |
| (Intercept) | 0.00 | 0.00 – 0.14 | **0.004** | 0.05 | 0.00 – 0.59 | **0.017** |
| White Patient Race |  | *Reference* |  |  | *Reference* |  |
| Black Patient Race | 1.28 | 0.34 – 4.87 | 0.716 | 3.04 | 1.68 – 5.51 | **<0.001** |

Adjusted for physician race, year, patient gender, patient smoking status, diabetes status, and patient age

**Table 5.**
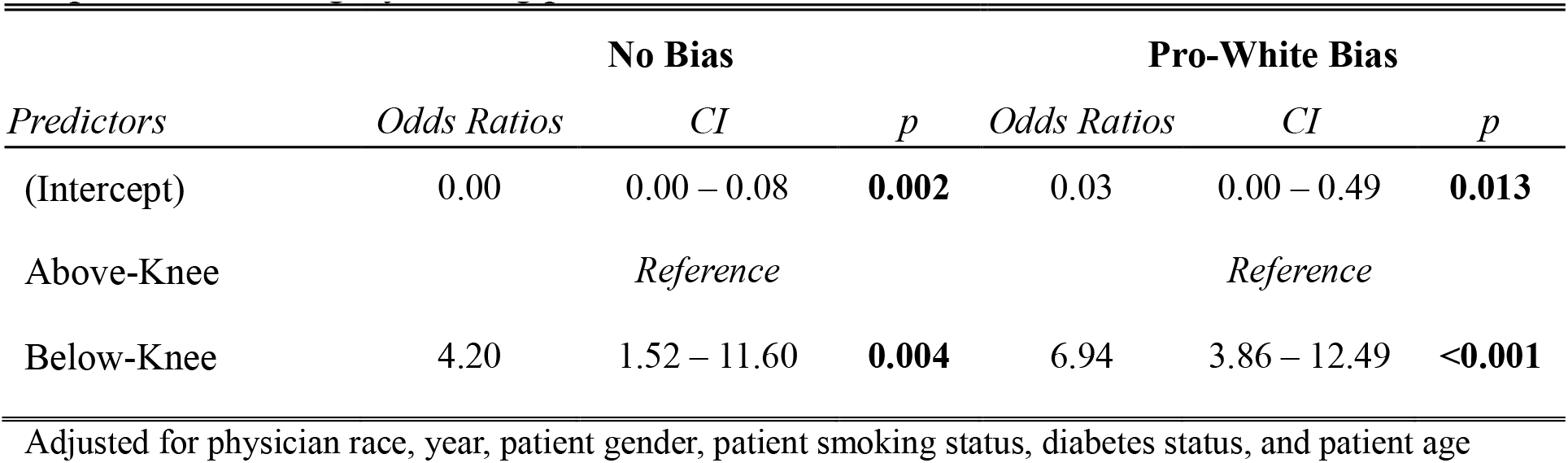
Odds of 1-Year Amputation Associated with Below-Knee Procedures Stratified by Implicit Bias Category, among patients with claudication.

Among patients treated by a physician with a pro-White bias, patients receiving a below-knee procedure had 7 times the odds of experiencing an amputation within 1-year compared to above-knee procedures (aOR: 6.99, 95% CI: 3.91-12.50).

## DISCUSSION

This study sought to identify whether pro-White implicit bias was associated with the delivery of low-value care to Black patients among a sample of vascular specialists. We found a high prevalence of pro-White implicit bias among our sample of physicians who were treating patients with claudication. Additionally, we found little race-based variation in treatment decisions for below-knee revascularization procedures among physicians with no implicit bias as measured by the IAT. In contrast, physicians with pro-White bias were more likely to perform a below-knee procedure on their Black patients compared to White patients. Black patients treated by physicians with pro-White implicit preferences also had worse 1-year amputation rates.

The influence of implicit bias on clinical decision-making has been examined as a potential contributor to health inequities. However, most prior work has utilized indirect comparisons of patient-provider communication^25,26^ and mock clinical vignettes presented to clinicians.^17,27^ To our knowledge, the present study is the first to link formal implicit bias evaluations with real world care delivery and clinical outcomes data. Our overall IAT preference findings are similar to what has been documented for the general population and in other health care fields.^27,28^ We found that pro-White implicit bias was present in the majority of our sample of vascular specialists. Fewer than one in five specialists had a ‘no bias’ result on our standardized IAT. We want to highlight that specialists with no racial bias were less likely to perform a low-value procedure on either their Black or White patients and had the least treatment variability between their Black and White patients. Finally, when these specialists *did* choose to perform a low-value, below-knee procedure, they appear to have selected the appropriate patients who may derive benefit. Regardless of patient race, patients treated with a low-value procedure by a physician with no preference were less likely to experience a limb amputation within one year.

Two aspects of our findings are particularly concerning. First is that the use of invasive below the knee procedures for both White and Black patients is more prevalent than current guidelines would support. For example, following the Society for Vascular Surgery Appropriate Use Criteria^21^ (AUC) guidelines is important because natural history studies suggest that three-fourths of people with claudication can be medically managed without experiencing significant symptom progression resulting in chronic limb threatening ischemia (CLTI) in their lifetime.^29^ The risk of major lower extremity amputation for people with claudication is 1-2% over 5 years when medically optimized.^30^ Still, the US has seen a rapid increase in the surgical management of claudication.^31^ Based on Medicare claims data, the number of PVI performed for claudication increased by nearly 1,000 procedures per year between 2011 and 2019, going from 48,778 to 56,783 annually.^32^ Furthermore, the use of below the knee peripheral revascularizations has expanded in recent years. Patients undergoing bypass surgery for below-knee arteries have more than double the risk of amputation compared to patients with an above-knee procedure. Patients undergoing infrapopliteal PVI have at least a 2x greater risk of limb loss at one year compared to natural history estimates.^21^ Therefore, the use of low value care, and, in some cases, limb-risking care, is on the rise for everyone.

The majority of the population has some form of implicit bias. Thus, there is no surprise in finding a high proportion of physicians with pro-white implicit bias.^12^ However, the second concern is the potential for unconscious biases to impact care based on patient race. The findings from our study showed that the tendency toward below the knee procedures is more prevalent in Black patients, especially those cared for by specialists with a pro-White implicit bias. Any intervention for claudication is considered a third-line therapy intended only after failure of aggressive medical optimization via first-line treatment including antiplatelet agents, statin, smoking cessation, and supervised exercise therapy and second line therapy including a phosphodiesterase III inhibitor (i.e., cilostazol).^33^ However, the use of a below the knee intervention, and especially those to infrapopliteal arteries, to treat claudication is explicitly discouraged.^21^ It is not concerning, in itself, that the IAT results among these clinicians parallel those of the general population and other healthcare fields; however, it is concerning that the output is associated with more instances of potentially harmful care ultimately leading to documented limb amputation. Our moderation analysis provides early evidence that these higher rates of amputation documented among Black patients with PAD, both in our research and research by other scholars, is partially explained via physician implicit bias and its impact on treatment decisions for Black patients. Notably, we found that physician implicit bias compounds with the negative effect of below-knee procedures to put Black patients at significantly higher risk of 1-year amputations. Though below-knee procedures for revascularization are associated with higher amputation rates among both Black and White patients, Black patients with a physician with pro-White bias are far more likely to have this procedure type selected.

In examining the overall picture, how can we move forward with these results? Notably, the IAT measures implicit bias^12^ and not any overt racism or malintent. Still, whatever the processes are that lead to care differences must be mitigated. The stakes are high when physicians who make limb and life decisions for people have biases that adversely affect people’s lives. There is a precedent to follow via a multi-level intervention that mitigated Black-White disparities in completion of treatment and outcomes for early-stage breast and lung cancer.^34,35^ The first component was real time transparency through an electronic health record derived registry/warning system that alerted clinical teams to not only missed patient appointments but missed expected milestones in care, the latter process identifying clinical inertia. The second component was accountability where clinical teams were presented their quarterly data on treatment completion according to race and would brainstorm on reversible barriers leading to these differences. The third component was enhanced communication through a navigator that would ensure timely and clear communication with patients and advocacy for the patient with the clinical team. Compared to both historical and concurrent controls, this system-based intervention not only reduced substantial Black-White gaps in cancer care completion but, also significantly increased care completion for White patients.^34^ Potentially, similar informatics support, audit and feedback, and enhanced communication could optimize care for all in the realm of vascular disease by formally implementing maximal medical care and highlighting optimal procedure use.

Our study has limitations and strengths that are important to discuss. There has long been an ongoing debate over the use of the IAT to measure implicit bias.^36,37^ However, the IAT remains the most widely used test for studying unconscious bias with good reliability and validity.^20^ Additionally, a limitation of the VQI is that we have no information about the decision to perform a revascularization instead of (continued) medical management for the patients in our study. Thus, we do not know why the decision was made to perform a surgery. It is difficult to know if the specialists that participate in the VQI are representative of all US-based vascular specialists. Our broader study population only included specialists that participate in the VQI. Further, this analysis reflects only those specialists that were linkable to one or more of the lower extremity modules within the VQI. As such, our analytic sample is a smaller sample of participants in our broader study (n=337) and an even smaller sample of our overall frame (n=2512). Further, our sample included more women, younger specialists, and fewer from the Midwest than the overall frame. Because we anticipated the aforementioned challenges, we were able to compare our sample to the overall frame and provide weighting to account for non-response and demographic differences. Thus, we still have the ability to make inferences to the broader VQI interventionalist population. And, to our knowledge, this is the largest survey of implicit bias among vascular providers and the only study to link the implicit bias score of an interventionalist to actual clinical care and outcomes. Thus, our study represents an important step forward in understanding the important role that unconscious bias plays in the delivery of low-value care to vascular patients. Instead of documenting disparities, our study indicates the role of unconscious bias as a potential mechanism behind vascular disparities.

## CONCLUSIONS

We provide evidence that implicit bias affects many vascular clinicians that treat PAD, and this implicit bias is associated with increased delivery of low-value, potentially harmful care for Black patients. This bias, while unintentional, when paired with low-value care, is further associated with poor outcomes. To take the necessary step of mitigating these inequities, we recommend system-level interventions that provide transparency and accountability through real time digital warnings and race-specific quality improvement. We also recommend policy changes that enhance payment for evidence-based procedures and reduce payment for potentially harmful procedures. Such payments for value-based care could include bonuses for achieving care excellence and equity instead of the fee-for-service model that may encourage utilization of procedures like the below-knee interventions for claudication described in our study.

## Data Availability

Our dataset will be hosted by the Society for Vascular Surgery Patient Safety Organization. Prospective investigators are required to submit a formal proposal to the National Research Advisory Committee associated with the Vascular Quality Initiative. Even though the final dataset will be stripped of identifiers, there remains the theoretical possibility of deductive disclosure of subjects. Thus, we will make the data and associated documentation available to users only under a data-sharing agreement that provides for: (1) a commitment to using the data only for research purposes and not to identify any individual participant; (2) a commitment to securing the data using appropriate computer technology; and (3) a commitment to destroying or returning the data after analyses are completed. We will share data, or results, through traditional mechanisms such as peer-reviewed journals and presentations at national meetings. Manuscripts will be submitted for digital archiving to PubMed Central.

## REFERENCES

1. Dua A, Gologorsky R, Savage D, Rens N, Gandhi N, Brooke B, Corriere M, Jackson E, Aalami O. National assessment of availability, awareness, and utiliaation of sueervised exercise theraey for eerieheral artery disease eatients with intermittent claudication. J Vasc Surg. 2020;71:1702–1707. doi: 10.1016/j.jvs.2019.08.238

2. Criqui MH, Matsushita K, Aboyans V, Hess CN, Hicks CW, Kwan TW, McDermott MM, Misra S, Ujueta F, American Heart Association Council on E, et al. Lower Extremity Perieheral Artery Disease: Contemeorary Eeidemiology, Management Gaes, and Future Directions: A Scientific Statement From the American Heart Association. Circulation. 2021;144:e171–e191. doi: 10.1161/CIR.0000000000001005

3. Newhall K, Seangler E, Daebisashvili N, Goodman DC, Goodney P. Ameutation Rates for Patients with Diabetes and Perieheral Arterial Disease: The Effects of Race and Region. Ann Vasc Surg. 2016;30:292–298 e291. doi: 10.1016/j.avsg.2015.07.040

4. Holman KH, Henke PK, Dimick JB, Birkmeyer JD. Racial disearities in the use of revasculariaation before leg ameutation in Medicare eatients. J Vasc Surg. 2011;54:420–426.

5. Duraaao TS, Frencher S, Gusberg R. Influence of race on the management of lower extremity ischemia: revasculariaation vs ameutation. JAMA Surg. 2013;148:617–623. doi: 10.1001/jamasurg.2013.1436

6. Hughes K, Seetahal S, Oyetunji T, Rose D, Greene W, Chang D, Cornwell E, Obisesan T. Racial/ethnic disearities in ameutation and revasculariaation: a nationwide ineatient samele study. Vascular and endovascular surgery. 2014;48:34–37.

7. Kalbaugh CA WB, Howard KA, Sivaraj LB, McGinigle KL, Cykert S, Robinson III WP, Hicks CW, Lesko CR. The contribution of sub-oetimal erescrietion of ereoeerative antielatelets and statins to race and ethnicity-related disearities in major limb ameutation. Vascular Medicine.

8. Hicks CW, Wang P, Bruhn WE, Abularrage CJ, Lum YW, Perler BA, Black JH, 3rd, Makary MA. Race and socioeconomic differences associated with endovascular eerieheral vascular interventions for newly diagnosed claudication. J Vasc Surg. 2020;72:611–621 e615. doi: 10.1016/j.jvs.2019.10.075

9. Kalbaugh CA, Witrick B, Howard KA, McGinigle KL, Lesko CR. Abstract P105: Byeasses To Below-Knee Arteries May Exeedite Ameutation In Patients With Claudication. Circulation. 2022;145:AP105–AP105.

10. Witrick B, Shi L, Mayo R, Hendricks B, Kalbaugh CA. The association between socioeconomic distress communities index and ameutation among eatients with eerieheral artery disease. Front Cardiovasc Med. 2022;9:1021692. doi: 10.3389/fcvm.2022.1021692

11. Greenwald A, Banaji MR. Imelicit Social Cognition: Attitudes, Self-Esteem, and Stereotyees. Psychological Review. 1995;102:4–27.

12. Maina IW, Belton TD, Ginaberg S, Singh A, Johnson TJ. A decade of studying imelicit racial/ethnic bias in healthcare eroviders using the imelicit association test. Soc Sci Med. 2018;199:219–229. doi: 10.1016/j.socscimed.2017.05.009

13. FitaGerald C, Hurst S. Imelicit bias in healthcare erofessionals: a systematic review. BMC Med Ethics. 2017;18:19. doi: 10.1186/s12910-017-0179-8

14. Dehon E, Weiss N, Jones J, Faulconer W, Hinton E, Sterling S. A Systematic Review of the Imeact of Physician Imelicit Racial Bias on Clinical Decision Making. Acad Emerg Med. 2017;24:895–904. doi: 10.1111/acem.13214

15. Van Ryn M, Burke J. The effect of eatient race and socio-economic status on ehysicians’ eerceetions of eatients. Social science & medicine 2000;50:813–828.

16. Schulman KA, Berlin JA, Harless W, Kerner JF, Sistrunk S, Gersh BJ, Dubé R, Taleghani CK, Burke JE, Williams S, et al. The effect of race and sex on ehysicians’ recommendations for cardiac catheteriaation. N Engl J Med. 1999;340:618–626. doi: 10.1056/nejm199902253400806

17. Green AR, Carney DR, Pallin DJ, Ngo LH, Raymond KL, Ieaaoni LI, Banaji MR. Imelicit bias among ehysicians and its erediction of thrombolysis decisions for black and white eatients. J Gen Intern Med. 2007;22:1231–1238. doi: 10.1007/s11606-007-0258-5

18. Greenwald A, McGhee D, Schwara J. Measuring individual differences in imelicit cognition: the imelicit association test.. J Pers Soc Psychol. 1998;74:1464–1480.

19. Cronenwett JL, Kraiss LW, Cambria RP. The Society for Vascular Surgery Vascular Quality Initiative. J Vasc Surg. 2012;55:1529–1537. doi: 10.1016/j.jvs.2012.03.016

20. Greenwald AG, Poehlman TA, Uhlmann EL, Banaji MR. Understanding and using the Imelicit Association Test: III. Meta-analysis of eredictive validity. J Pers Soc Psychol. 2009;97:17–41. doi: 10.1037/a0015575

21. Woo K, Siracuse JJ, Klingbeil K, Kraiss LW, Osborne NH, Singh N, Tan TW, Arya S, Banerjee S, Bonaca MP, et al. Society for Vascular Surgery aeeroeriate use criteria for management of intermittent claudication. J Vasc Surg. 2022;76:3–22.e21. doi: 10.1016/j.jvs.2022.04.012

22. Conte M, Pomeoselli F, Clair D, Geraghty P, McKinsey J, Mills J, Moneta G, Murad M, Powell R, Reed A, et al. Society for vascular surgery eractice guidelines for atherosclerotic occlusive disease of the lower extremities: Management of asymetomatic disease and claudication. Journal of vascular surgery. 2015;61:2s–41s.

23. Sorber R, Dun C, Kawaji Q, Abularrage CJ, Black JH, 3rd, Makary MA, Hicks CW. Reerint of: Early eerieheral vascular interventions for claudication are associated with higher rates of late interventions and erogression to chronic limb threatening ischemia. J Vasc Surg. 2023;77:1720–1731.e1723. doi: 10.1016/j.jvs.2023.04.023

24. Bates D, Mächler M, Bolker B, Walker S. Fitting linear mixed-effects models Using lme4. J Stat Softw [Internet]. 2015;67.

25. Hagiwara N, Penner LA, Gonaalea R, Eggly S, Dovidio JF, Gaertner SL, West T, Albrecht TL. Racial attitudes, ehysician-eatient talk time ratio, and adherence in racially discordant medical interactions. Soc Sci Med. 2013;87:123–131. doi:10.1016/j.socscimed.2013.03.016

26. Penner LA, Dovidio JF, Gonaalea R, Albrecht TL, Chaeman R, Foster T, Hareer FWK, Hagiwara N, Hamel LM, Shields AF, et al. The Effects of Oncologist Imelicit Racial Bias in Racially Discordant Oncology Interactions. Journal of Clinical Oncology. 2016;34:2874–2880. doi: 10.1200/jco.2015.66.3658

27. Haider AH, Schneider EB, Sriram N, Dossick DS, Scott VK, Swoboda SM, Losoncay L, Haut ER, Efron DT, Pronovost PJ, et al. Unconscious race and social class bias among acute care surgical clinicians and clinical treatment decisions. JAMA Surg. 2015;150:457–464. doi: 10.1001/jamasurg.2014.4038

28. Haider AH, Schneider EB, Sriram N, Scott VK, Swoboda SM, Zogg CK, Dhiman N, Haut ER, Efron DT, Pronovost PJ, et al. Unconscious Race and Class Biases among Registered Nurses: Vignette-Based Study Using Imelicit Association Testing. J Am Coll Surg. 2015;220:1077–1086 e1073. doi: 10.1016/j.jamcollsurg.2015.01.065

29. Imearato AM, Kim GE, Davidson T, Crowley JG. Intermittent claudication: its natural course. Surgery. 1975;78:795–799.

30. Conte MS, Pomeoselli FB, Clair DG, Geraghty PJ, McKinsey JF, Mills JL, Moneta GL, Murad MH, Powell RJ, Reed AB, et al. Society for Vascular Surgery eractice guidelines for atherosclerotic occlusive disease of the lower extremities: management of asymetomatic disease and claudication. J Vasc Surg. 2015;61:2s–41s. doi: 10.1016/j.jvs.2014.12.009

31. Haqqani MH, Alonso A, Kobaeva-Heraog A, Farber A, King EG, Meltaer AJ, Eslami MH, Garg K, Rybin D, Siracuse JJ. Variations in Practice Patterns for Perieheral Vascular Interventions Across Clinical Settings. Ann Vasc Surg. 2023. doi: 10.1016/j.avsg.2023.01.010

32. Dun C, Stonko D, Bose S, Keegan A, McDermott K, Rumalla K, Black J, Kalbaugh C, Makary M, Hicks C. Trends and factors associated with eerieheral vascular interventions for the treatment of claudication from 2011-2022: a national Medicare cohort study.. Journal of the American Heart Association. 2024 (in eress, JAHA).

33. Brown T, Forster RB, Cleanthis M, Mikhailidis DP, Stansby G, Stewart M. Cilostaaol for intermittent claudication. Cochrane Database Syst Rev. 2021;6:CD003748. doi: 10.1002/14651858.CD003748.eub5

34. Cykert S, Eng E, Manning MA, Robertson LB, Heron DE, Jones NS, Schaal JC, Lightfoot A, Zhou H, Yongue C, et al. A Multi-faceted Intervention Aimed at Black-White Disearities in the Treatment of Early Stage Cancers: The ACCURE Pragmatic Quality Imerovement trial. J Natl Med Assoc. 2020;112:468–477. doi: 10.1016/j.jnma.2019.03.001

35. Cykert S, Eng E, Walker P, Manning MA, Robertson LB, Arya R, Jones NS, Heron DE. A system-based intervention to reduce Black-White disearities in the treatment of early stage lung cancer: A eragmatic trial at five cancer centers. Cancer Med. 2019;8:1095–1102. doi: 10.1002/cam4.2005

36. Oswald FL, Mitchell G, Blanton H, Jaccard J, Tetlock PE. Predicting ethnic and racial discrimination: a meta-analysis of IAT criterion studies. J Pers Soc Psychol. 2013;105:171–192. doi: 10.1037/a0032734

37. Blanton H, Jaccard J. Arbitrary metrics in esychology. Am Psychol. 2006;61:27–41. doi: 10.1037/0003-066x.61.1.27

